# Initial real world evidence for lower viral load of individuals who have been vaccinated by BNT162b2

**DOI:** 10.1101/2021.02.08.21251329

**Authors:** Ella Petter, Orna Mor, Neta Zuckerman, Danit Oz-Levi, Asaf Younger, Dvir Aran, Yaniv Erlich

## Abstract

One of the key questions regarding COVID19 vaccines is whether they can reduce viral shedding. To date, Israel vaccinated substantial parts of the adult population, which enables extracting real world signals. The vaccination rollout started on Dec 20th 2020, utilized mainly the BNT162b2 vaccine, and focused on individuals who are 60 years or older. By now, more than 75% of the individuals of this age group have been at least 14 days after the first dose, compared to 25% of the individuals between ages 40-60 years old. Here, we traced the Ct value distribution of 16,297 positive qPCR tests in our lab between Dec 1st to Jan 31st that came from these two age groups. As we do not have access to the vaccine status of each test, our hypothesis was that if vaccines reduce viral load, we should see a difference in the Ct values between these two age groups in late January but not before. Consistent with this hypothesis, until Jan 15th, we did not find any statistically significant differences in the average Ct value between the groups. In stark contrast, our results in the last two weeks of January show a significant weakening in the average Ct value of 60+ individuals to the 40-60 group. To further corroborate these results, we also used a series nested linear models to explain the Ct values of the positive tests. This analysis favored a model that included an interaction between age and the late January time period, consistent with the effect of vaccination. We then used demographic data and the daily vaccination rates to estimate the effect of vaccination on viral load reduction. Our estimate suggests that vaccination reduces the viral load by 1.6x to 20x in individuals who are positive for SARS-CoV-2. This estimate might improve after more individuals receive the second dose. Taken together, our findings indicate vaccination is not only important for individual’s protection but can reduce transmission.

## Introduction

Vaccines are considered the main exit strategy from the COVID19 pandemic. Large scale phase 3 Randomized Controlled Trials (RCT) have shown that mRNA-based vaccines can substantially reduce symptomatic cases with estimated efficacy of 95% (*1, 2*). However, a key question is the ability of vaccines to also cut the transmission of the virus, which is necessary to reduce the risk of emerging strains and ultimately achieving herd immunity (*3*). Previous RCT provided limited data on vaccine efficacy with respect to transmission (*4*) and no real world data has been reported.

Since December 20th 2020, Israel has rolled a massive COVID19 vaccination program mainly using the BioNTech/Pfizer BNT162b2, which consists of two shots separated by 21 days. The program started by immunization of high risk individuals, which were defined mainly as 60+ years old, medical personnel, workers at nursing homes, and individuals with comorbidities. This phase lasted until January 21st afterwhich the age restriction limit was gradually lowered. By Feb 4th, every Israeli citizen above 16 year old is eligible for the vaccine unless they have a medical condition that prevents that. As of this date, 77% of the individuals at ages 60-80 received at least one dose and nearly 90% of 80+ individuals received at least one dose.

During the same period of time, Israel experienced a “third wave” of the pandemic. This wave was more brutal than the previous two and resulted in more than 1,400 COVID19 related deaths during January, a record number. In addition, sequencing data of 460 random positive samples by the Ministry of Health’s Central Virology lab indicated the emergence of SARS-CoV-2 variants of concern (B.1.1.7 and B.1.351) during the past two months (**Table S1**). Specifically, at late December, only 5% of the samples had a variant of concern, while on Jan 18th, about 50% exhibited these variants. From all 57 samples with variants of concern, only 5 were B.1.351 and the rest were B.1.1.7.

Looking for signs of transmission reduction by vaccines, we sought to analyze the Ct value of positive qPCR tests collected between December 1st to February 1st 2021 at the MyHeritage lab, the largest COVID19 testing lab in Israel. The lab typically processes between 10,000 to 20,000 samples a day and has processed over 2.7 million samples since June 2020, approximately 30% of the total COVID-19 qPCR tests in Israel. For all tests, we use a single assay, the BGI SARS-CoV-2 RT-PCR testing kit. This assay detects the Orf1_ab gene of the SARS-CoV-2 virus on the FAM fluorescent channel and the human beta-actin on the VIC fluorescent channel. This provides consistency with respect to the results and allows comparison between data points.

Importantly, the lab does not have individual-level health data on any of the testees besides very basic demographic infomration such as age and sex. Thus, we were unable to trace the vaccination status of each person. Instead, we used the vaccination criteria as a natural experiment. As other studies (*5*), our expectation was that during January, testees over the age of 60 consists of people who have been vaccinated in increasing numbers, whereas other younger individuals mainly consist of individuals who did not receive the vaccine. This way, we can approximate the effect of the vaccine on the Ct values despite the significant changes in the dynamic of the pandemic in Israel and the emergence of the variant of concerns.

## Results

To this end, we traced the Ct value distribution of positive tests stratified the results based on the age group of the individuals between December 1st, 2020 to January 30th (**Figure 1**). The tests were called based on our SOP (standard operating procedure) which requires Ct<36 to define a test as a definite positive given that a reaction curve is present and Ct<33 for the beta-actin human control. In total, we had 16,297 positive tests and we traced the average Ct values stratified to two age groups: individuals over 60 (5,317 positive tests) and individuals between 40 to 60 years old (10,980 positive tests). Consistent with previous studies (*6*), we did not observe substantially higher viral load due to the emergence of B.1.1.7 in Israel.

**Figure 1:**
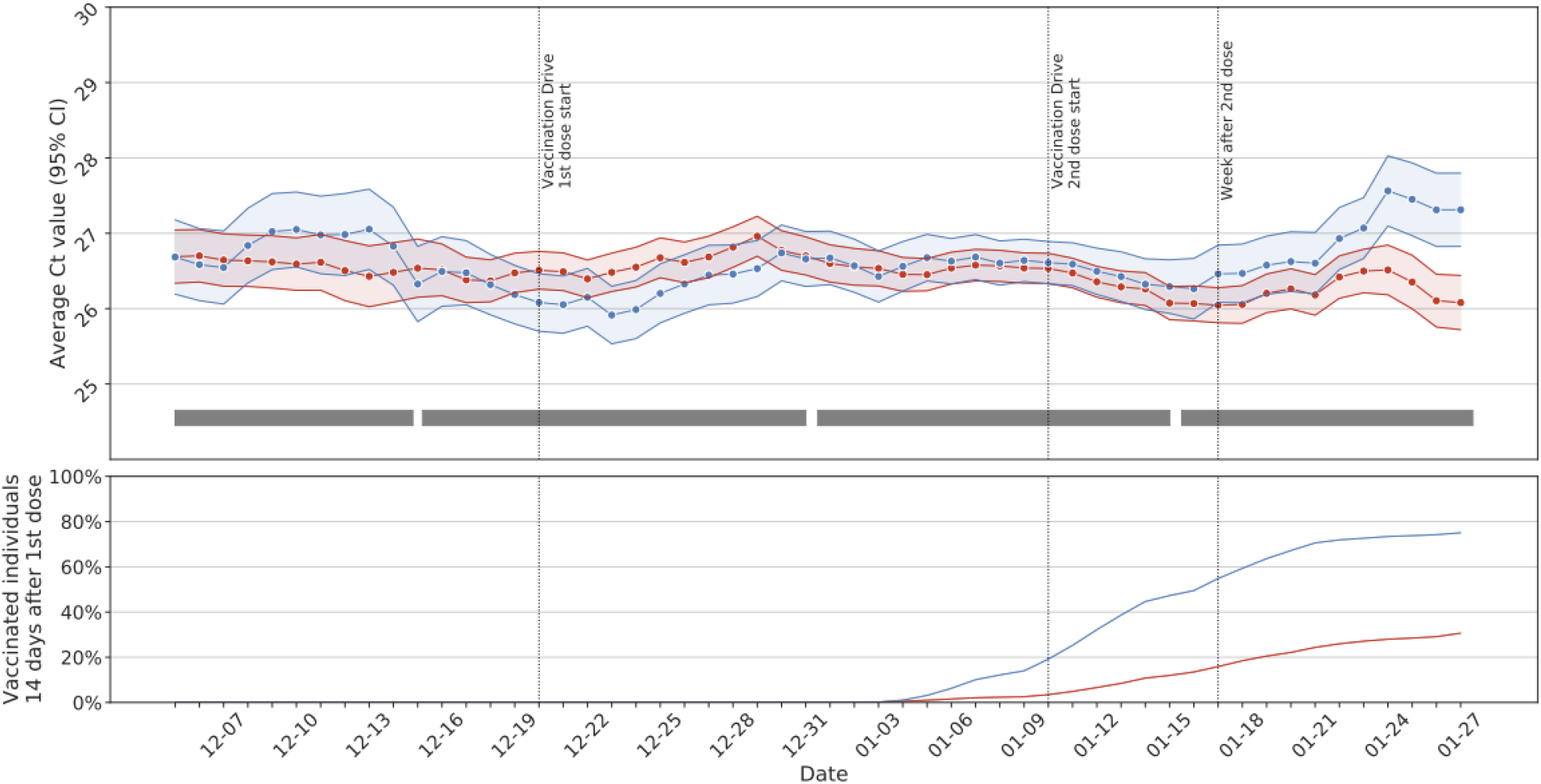
The Ct value distribution as a function of date. Blue: 60+ individuals. Red: Individuals between the ages of 40-60yrs. Upper: the average Ct value of positive tests per age group smoothed using a 7 day rolling window. The shadowed area shows the 95% confidence interval. The gray rectangles represent the four comparison periods in **Figure 2**. The nationwide percentage of individuals who are 14 days after receiving their first dose [data processed from the Ministry of Health dashboard].

Our results show small but significant differences in Ct value patterns between the age groups before and after vaccinations. Before making any analysis, we selected January 15th as our cutoff to evaluate the effect of vaccination. As such, our results avoid post-hoc issues. We then compared the average Ct value in the 60+ individuals vs. the individuals between 40-60 years old across four periods: (i) Dec 1st to Dec 15th (ii) Dec 16th to Dec 31st (iii) Jan 1st to Jan 15th (iv) Jan 15th to Jan 30th. We did not find any significant difference in the average Ct values between the groups in the first three time periods. Importantly, the fourth time period exhibited a statistically significant difference (t-test, bonferroni-corrected p<7×10^−6^). The 60+ group had Ct values that were weaker (i.e, numerically higher) by 0.82 (95% CI: 0.66-0.97) cycles compared to the younger group (**Figure 2**).

**Figure 2:**
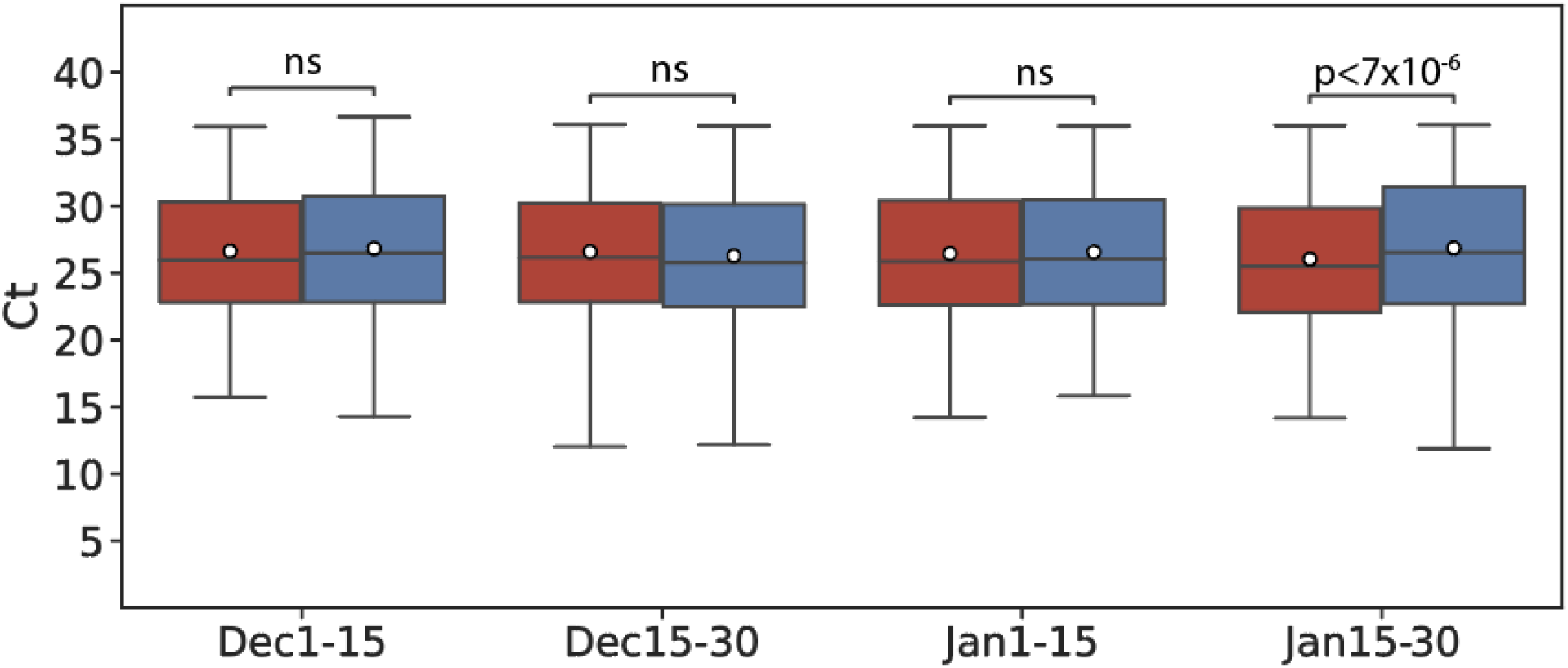
The Ct values for the two age groups as a function of time. Blue: 60+ individuals. Red: Individuals between age 40-60yrs. The first three time periods did not exhibit any statistically significant differences between the average Ct values. After Jan 15th, the individuals over age 60 showed a statistically significant weaker Ct value compared to individuals between age 40 to 60.

To further corroborate our findings, we evaluated several nested linear models to explain the data (**Table 1**). In Model I, we regressed the Ct value of each positive test on the the sex and the age group (40-60 or 60+). This model can be thought of as a basic demographic model to explain the Ct value distribution. Model II had the independent variables of Model I and also included an indicator variable that was set to 1 if the test was conducted in the last two weeks of January. This model can be thought of as a demographic model that allows a temporal change in the Ct values in late January for all age groups (for instance due to an emerging strain). Model III had the independent variables of Model II and also included an interaction term between the age group and whether the test was conducted in the past two weeks of January. This model includes all demographic variables and a temporal change that is age category specific and approximates the effect of higher vaccination rates for 60+ individuals.

**Table 1:** model selection. The circles represent the inclusion of various variables (gray: non-significant; black: significant after Benefroni correction)

|  | Model I | Model II | Model III |
| --- | --- | --- | --- |
| **Independent variables** |  |  |  |
| Sex | ◉ | ◉ | ◉ |
| Age group (40-60 individuals vs. 60+ individuals) | ◉ | ◉ | ◉<br>p<0.01 |
| Late January |  | ◉<br>p<5x10<sup>-4</sup> | ◉<br>p<2x10<sup>-5</sup> |
| Late January-by-Age group |  |  | ◉<br>p<0.026 |
| **ΔAIC vs. Model I** | n/a | -12 | -17 |

Our model comparison preferred Model III that is more consistent with vaccination over the other models. In Model I, the two independent variables were not statistically significant, arguing against a simple model that relies on age group and sex. In Model II, the late January variable was statistically significant (t-test; bonferroni-corrected p<0.0005). In Model III, the interaction term was significant (t-test; bonferroni-corrected p<0.026) together with age group (t-test; bonferroni-corrected p<0.01) and also the late January period (t-test; p<2×10^−5^) but not the late January variable. The **Δ** AIC between the Model 2 to Model 3 was 5.3, favoring model III.

Encouraged by these results, we sought to estimate the effect of vaccination on viral load. Our results indicated an average Ct value difference of 0.82 in late January between 60+ groups and the 40-60 group. However, the reduction in viral load among vaccinated individuals can be more substantial than reflected by our results. First, as vaccines reduce the rate of active cases, the rate of positive 60+ individuals who are vaccinated is smaller than the rate of the population. Second, by Jan 31st, about 30% of the 40-60 group were 14 days after the first dose compared to ∼75% of the 60+ group. This can reduce the difference in the Ct values between the groups.

To this end, we constructed a model to estimate the effect of vaccination on viral load in positive individuals. Let Δdenote the effect of the vaccine on the Ct value, which is assumed to be identical in time and age groups. Our model further assumes that each day, we receive two types of positive samples, from vaccinated individuals and unvaccinated individuals. Let *α*_*g*_ (*i*) denote the fraction of positive samples of vaccinated people for the age group, *g*, at the i-th day. We estimate 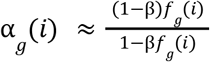, where *f*_*g*_(*i*)denoted the fraction of individuals in age group *g* at the i-th day who are 14 days after the first dose and as the effectiveness of the vaccinate to reduce the number of positive individuals compared to unvaccinated. The daily estimate of Δ is 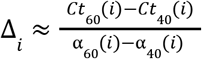, where *Ct*_*g*_(*i*)denotes the average Ct value for the age group g. We then approximate Δas the weighted average of the daily estimates using the inverse variance weighting.

We evaluated this model by estimating the daily Δ_*i*_ between Jan 15th to Jan 30th (**Figure 3**). In our previous studies we found that the effectiveness in reducing the number of <u>positives</u> grows from β = 0. 4 to β = 0.8 between 14 to day 21 days post the first dose (*7*). To accommodate this heterogeneity, we ran the model using these two effectiveness levels, where the former is more conservative and the latter is more permissive with respect to the estimates of viral load reduction.

**Figure 3:**
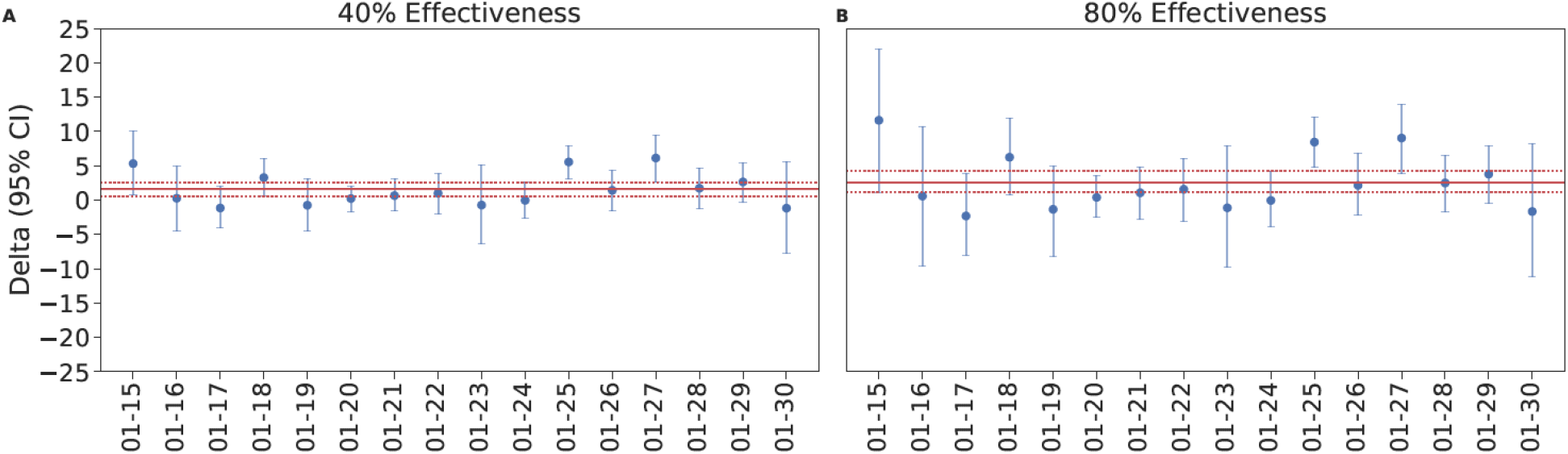
Estimating the Ct value weakening by vaccination. In both figures, the blue circles denote the expected Δand the bars denote the 95% confidence interval calculated by bootstrap. The solid red lines are the maximum likelihood estimator for Δin each model. The dotted red lines are the 95% confidence intervals for the estimate (A) The model using β = 0. 4 (B) The model using β = 0.8

Our results show that the Ct value weakening of positives who are vaccinated is 1.61 cycles for the permissive model (95% CI: 0.72-2.67; bootstrapping using 10,000 rounds) to 2.61 in the more restrictive model (95% CI: 1.19-4.29; bootstrapping using 10,000 rounds). Taken together, our models predict that the expected viral load of vaccinated individuals who are positive to SARS-CoV-2 to be between 1.6x to 20x fold lower than unvaccinated individuals.

## Discussion

We traced the Ct value of samples in our lab to estimate the effect of vaccination on viral load. Despite using data without health information, we approximated the effects by utilizing the vaccination patterns in Israel between age groups. Our results showed a statistically significant difference in the average Ct value in late January but not before between 60+ individuals, who were the first to be vaccinated, and 40-60 individuals that were vaccinated in lower rates by this time point. We then employed a series of nested linear models to explain this pattern by demographic patterns, an emerging strain by the end of January, or by vaccination differences between the two ago groups. Our model comparison favored the vaccination model to explain the difference in the average Ct value, providing further evidence. Finally, we provided an initial estimate of the Ct value differences due to vaccination using a basic model that takes into account vaccination rates of each group and the presumed effectiveness of the vaccine in reducing the number of positives. Our results predict that positive vaccinated individuals are expected to have a lower viral load that is proportional to 0.72-4.29 cycles.

Importantly, our analysis has multiple caveats. First, we assumed that vaccination has a homogenous effect at all age groups. However, lower efficacy in older individuals means that our results probably underestimate the effect of vaccination on viral load. Second, we did not take into account the effect of the second dose. Again, this can further improve our estimated effects for viral load reductions. Third, we assumed that the emerging variants distribute equally among age groups. We believe that this assumption is a valid approximation since by mid January 50% of the thousands of daily cases had this strain, meaning that these variants were widely distributed in Israel by late January and that previous studies did not find differences in viral load between B.1.1.7 and wild type SARS-CoV-2 (*6*). Fourth, it has been estimated that 33% of all cases in Israel are due to household transmission. As about 50% of 60+ individuals live in a household of only two individuals and are likely to be both vaccinated, some of the events in our data must reflect the transmission from a vaccinated person to another vaccinated person. If the viral load is correlated between individuals, our analysis may overestimate to some extent the viral load reduction by vaccination. However, we believe that this confounder is relatively mild. Finally, our prediction relies on several demographic and effectiveness assumptions.

Despite these shortcomings, our estimates are encouraging. Previous studies have shown that viral load has been associated with transmission rates (*8, 9*) and COVID-19 disease severity (*10*). Therefore, our results indicate that while vaccinated individuals might be positive, they have lower viral load and therefore can be less infective and might also experience a milder disease.

Our findings highlight that vaccination does not only protect the individual who receives it but is likely to reduce viral shedding and therefore transmission in the population. As such, they point towards the value of investing efforts to include younger segments of the population in the vaccination efforts. At least in the Israeli population, these segments exhibit high rates of vaccine hesitancy. We posit that providing evidence that vaccines can reduce transmission and therefore protect older family members of these individuals can be an important tool to encourage young individuals to be vaccinated.

## Data Availability

Summary statistics will be uploaded to a public Github. Further public data is available here: https://data.gov.il/dataset/covid-19

## Acknowledgement

We thank Eran Segal from Weizmann Institute for useful discussions and the MyHeritage Lab employees for their work in collecting these data.

## Supplemental Material

**Table S1:** the presence of variants of concerns (B.1.1.7 or B.1.351) in randomly ascertained samples from the MyHeritage lab based on Illumina sequencing by the Ministry of Health Central Virology Lab.

| Date | Variant | Samples | Ratio(%) |
| --- | --- | --- | --- |
| 2020-12-27 | 8.0 | 167 | 4.8 |
| 2021-01-03 | 24.0 | 241 | 10.0 |
| 2021-01-14 | 4.0 | 8 | 50.0 |
| 2021-01-18 | 21.0 | 44 | 47.7 |

