## Supplementary material for "Initial real world evidence for lower viral load of individuals who have been vaccinated by BNT162b2": Methods

Samples

Human samples were obtained by nasopharyngeal swabbing. Sampled swabs were immediately kept in sterile vials containing universal transport medium (UTM). The vials were stored in coolers during transportation to the lab and refrigerated at 4˚C in the lab until processing.

Standard protocol (including RNA extraction)

Samples undergo three main steps of processing in our lab in our standard protocol:

(a) Heat inactivation: the samples are exposed to 70˚C for 45 minutes in an oven.

(b) RNA extraction: we use the MGIEasy RNA extraction kit that is manufactured by BGI's subsidiary company MGI. Briefly, 200ul of the sample is plated into a 1.3ml deep well plate. The RNA extraction is a semi-automated process that uses MGISP-960 robots that take 160ul and eventually produce 30ul of extracted genomic material from each sample, using magnetic beads.

(c) RT-qPCR: we use the BGI SARS-CoV-2 qPCR kit that detects Orf1_ab and the human beta-actin gene on the FAM and VIC channels, respectively. First, 10ul of RNA-extracted sample is added into 20ul of mix from the qPCR kit that contains the qPCR primers, probes, and nucleotides. Then, the qPCR program is as follows:

1. Reverse Transcription: 50˚C for 20 min.
2. Denaturation: 95˚C for 10 min.
3. Cycling: 45 cycles of 95˚C for 15 sec followed by 60˚C for 30 sec and fluorescent quantification.
4. Cooling: 37˚C for 10 sec.

The Ct value is calculated automatically by a qPCR instrument using the maximal second derivative. Analysis is then conducted by the lab staff using the Ct values of FAM and VIC and their respective graphs. Our lab currently uses Roche LightCycler 480 Instrument II and ThemoFisher ABI QuantStudio 5 qPCR instruments.

Thresholds

We used the following thresholds for classifying the samples based on our approved Standard Operating Procedure (SOP). If not stated otherwise, positives were defined as samples with a FAM Ct value of 15 to 36. Weak positives were defined as samples with a FAM Ct value of 36 to 38.5. Negatives were defined as everything else. FAM is the channel that detects the SARS-CoV-2 Orf1_ab gene.

**Epidemiological data**

Data about vaccination was taken from the Israeli Ministry of Health website:
https://data.gov.il/dataset/covid-19/resource/57410611-936c-49a6-ac3c-838171055b1f

Data about the ages of individuals was taken from the 2019 table of the Israeli Census:
https://www.cbs.gov.il/he/publications/LochutTlushim/2020/%D7%90%D7%95%D7%9B%D7%9C%D7%95%D7%A1%D7%99%D7%99%D7%942019-2000.xlsx

**Programs**

The analysis was conducted in Python3 using Pandas and Numpy, and partially in R. The statistical tests are written in the text. The code is available upon request from the first author.
